# INFEKTA: A General Agent-based Model for Transmission of Infectious Diseases: Studying the COVID-19 Propagation in Bogotá - Colombia

**DOI:** 10.1101/2020.04.06.20056119

**Authors:** Jonatan Gomez, Jeisson Prieto, Elizabeth Leon, Arles Rodríguez

## Abstract

The transmission dynamics of the coronavirus - COVID-19-have challenged humankind at almost every level. Currently, research groups around the globe are trying to figure out such transmission dynamics using different scientific and technological approaches. One of those is by using mathematical and computational models like the compartmental model or the agent-based models. In this paper, a general agent-based model, called INFEKTA, that combines the transmission dynamics of an infectious disease with agents (individuals) that can move on a complex network of accessible places defined over a Euclidean space representing a real town or city is proposed. The applicability of INFEKTA is shown by modeling the transmission dynamics of the COVID-19 in Bogotá city, the capital of Colombia.

## Introduction

Infectious diseases have a substantial impact on public health, health care, macroeconomics, and society. The availability of options to control and prevent the emergence, expansion or resurgence of pathogens warrants continuous evaluation using different methods. Mathematical models allow characterizing both the behavior and the emergent properties of biological systems, such as the transmission of infectious disease. [1–3]. Many biological systems have been modeled in terms of complexity since their collective behavior cannot be simply inferred from the understanding of their components [4, 5].

Complex systems are computational approaches that make use of computer-based algorithms to model dynamic interactions between individuals agents (e.g. persons, cells) or groups and their properties, within, and across levels of influence [6, 7]. In general, agent-based modeling (ABM) can be used for testing theories about underlying interaction mechanics among the system’s components and their resulting dynamics. It can be done by relaxing assumptions and/or altering the interaction mechanisms at the individual agent level. ABMs can increase our understanding of the mechanisms of complex dynamic systems, and the results of the simulations may be used for predicting future scenarios [8].

In this paper, we introduce an ABM, called INFEKTA (Esperanto word for infectious), for modeling the transmission of infectious disease, applied to the coronavirus COVID-19. INFEKTA models the disease transition at the person level and takes into consideration individual infection disease incubation periods and evolution, medical preconditions, age, daily routines (movements from house to destination places and back, including transportation medium if required), and enforced social separation policies.

### Complex Systems Approaches for Epidemic Models

The complex system model approach considers a system as a large number of entities (equally complex systems that have autonomous strategies and behaviors) that interact with each other in local and non-trivial ways [9–11]. This approach provides a conceptual structure (a multi-level complex network [12]) that allows characterizing the interrelation and interaction between elements of a system and between the system and its environment [13]. In this way, a system is composed of sub-systems of second order, which in turn may be composed of subsystems of the third-order [14]. Transmission dynamics of infectious diseases are not traditionally modeled at the individual level, but at the population-level with a compartmental model. However, some recent research use agent-based modeling for doing that [15].

### Compartmental Model

A compartmental model tracks changes in compartments without specifying which individuals are involved [16] and typically reflects health states relevant for transmission (e.g., susceptible, exposed, infectious and recovered). Basically, these kinds of models represent epidemics of the communicable disease using a population-based, non-spatial approach. The conceptual framework for this approach is rooted in the general population model which divides a population into different population compartments [17]. Compartmentalization typically reflects health states relevant for transmission (e.g., susceptible, exposed, infectious and recovered, in short **SEIR**), though more partitioning is possible according to age and/or other relevant host characteristics. Heterogeneous and temporal behavior is modeled through the incorporation of relevant time-dependent social mixing, community structures and seasonality, relevant for infectious disease dynamics [18, 19]. Process dynamics are captured in transition rates, representing the rate by which an average individual transitions between compartments.

### Agent-based models for infectious disease

Agent-based models (ABMs) are a type of computer simulation for the creation, disappearance, and movement of a finite collection of interacting individuals or agents with unique attributes regarding spatial location, physiological traits and/or social behavior [20–22]. ABMs work bottom-up, with population-level behavior emerging from the interactions between autonomous individuals and their environment [22, 23]. They allow the history of every individual to be tracked and network structures to be explicitly represented.

In general, ABMs allow: i) To introduce local interaction rules at the individual level, which closely coincide with physical and social interaction rules; (ii) To include behaviors that may be randomized at the observational level, but can be deterministic from a mathematical point of view; (iii) To incorporate a modular structure and to add information through new types of individuals or by modifying current rules; and (iv) To observe systems dynamic that could not be inferred from the examination of the rules of particular individuals [8].

Because of the complex nature of interactions among individuals, an ABM may not provide an understanding of the underlying mechanisms of the complex system. However, changes in the local rules repertoire of the individuals can be introduced into the ABM to observe changes in the complex system [24].

When ABM is used for epidemic modeling, infectious disease transmission dynamics is expected to emerge from the interaction between local interactions between the individuals. In this way, each virtual individual includes its health state relevant for the transmission of the infectious disease (severity and time in it), a kind of individual SEIR (or more complex) model. Health state transmission rates are usually approximated from the rates obtained by a compartmental model but are used at the individual level, i.e., when individuals interact with each other.

### INFEKTA Agent-Based Model

Our agent-based model of infectious disease propagation, called INFEKTA, consists of five-layer components: **i) Space:** virtual space where individuals move and interact; **ii) Time:** virtual time used by individuals for moving and interacting; **iii) Individuals:** virtual persons being simulated, **iv) Infectious disease dynamics:** Specific disease parameters that modeled how an individual can get infected and how the infectious disease will evolve; and **v) Social separation policy:** Set of rules defined for restricting both access to places and mobility of individuals.

#### Space

The virtual space (for a city or town being studied) is a Euclidean complex network [12]: Nodes are places (located in some position of the 2D Euclidean space) where individuals can be at some simulation time and edges are routes (straight lines) connecting two neighbor places.

#### Place (Node)

A place may be of three kinds: home (where individuals live), public transportation station (PTS), and interest place (IP) i.e., school, workplace, market, and transportation terminal. IPs and PTSs are defined in terms of capacity (maximum number of individuals that can be at some simulation step time). IPs and PTSs may be restricted, during some period, to some or all individuals. Place restriction is established according to the social separation rule that is enforced during such a period.

#### Neighbor (Edge)

A PTS is a neighbor to another according to the public transportation system of the city or town being studied. Homes and IPs are considered neighbors to its closest^1^ PTS. No home is neighbor to any other home neither an IP is neighbor of any other IP. Finally, a home and an IP are considered neighbors if they are neighbors of the same PTS.

### Time

Virtual time is defined in INFEKTA at two resolution levels: days for modeling the transmission dynamics of the infectious disease, and hours for modeling the moving and interaction of individuals. Therefore, if an individual gets infected more than once during the same day, INFEKTA considers all of them as a single infection event. Any individual movement is carried on the same one hour, it was started, regardless of the traveled Euclidean distance neither the length of the path (number of edges in the complex network).

### Individuals

A virtual individual in INFEKTA is defined in terms of his/her demographic, mobility and infectious disease state information.

#### Demographics

The demographic information of a virtual individual consists of: **i) Age** of the individual; **ii) Gender** of the individual *female* o *male*; **iii) Location** of the individual at the current time step; **iv) Home** of the individual, **v) Impact level of medical preconditions** on the infectious disease state if the individual is infected, and **vi) IP interest** of going to certain type of IPs.

#### Mobility

The ability of an individual to move through space (we use the graph defining the space for determining the route as proposed in [25, 26]). Each individual has a **mobility plan for every day**, plan that is carried on according to the enforced social separation policy and her/his infectious disease state.

The mobility plan is modeled in INFEKTA as a collection of simple movement plans to have **i) Policy**: social separation policy required for carrying on the mobility plan; **ii) Type**: may be *mandatory*, i.e., must go to the defined interest place) or *optional*, i.e., any place according to individual’s preferences; **iii) Day**: day of the week the plan is carried on, maybe *every, week, weekend, Monday*, …, *Sunday* ; **iv) Going Hour**; **v) Duration** in hours for coming back to home; and **vi) Place**: if plan type is mandatory, it is a specific place, otherwise it is an IP selected by the individual according to his/her IP preferences.

### Infectious Disease Dynamic

Any individual can potentially be in one of seven different infectious disease states or health states: Immune (*M*), Susceptible (*S*), Exposed (*E*), Seriously-Infected (*I*_*S*_), Critically-Infected (*I*_*C*_), Recovered (*R*), and Dead (*D*). As can be noticed, we just adapt the terminology from the compartmental models in epidemiology – namely, from the SEIR (Susceptible-Exposed-Infectious-Recovered) model. In INFEKTA, the infectious state of the SEIR model is divided into exposed, seriously-infected and critically infected in order to capture how age, gender, IP preferences, medical preconditions (co-morbidity), and social separation policies can impact the evolution of the infectious disease in an individual. INFEKTA introduces both the *M* state since some individuals are naturally immune to or can become immune to (after recovering) to certain infectious diseases and the Dead (*D*) state to distinguish between recovered and dead individuals. Figure 1 shows the transition dynamics of the infectious disease at the individual level in INFEKTA.

**Fig 1.**
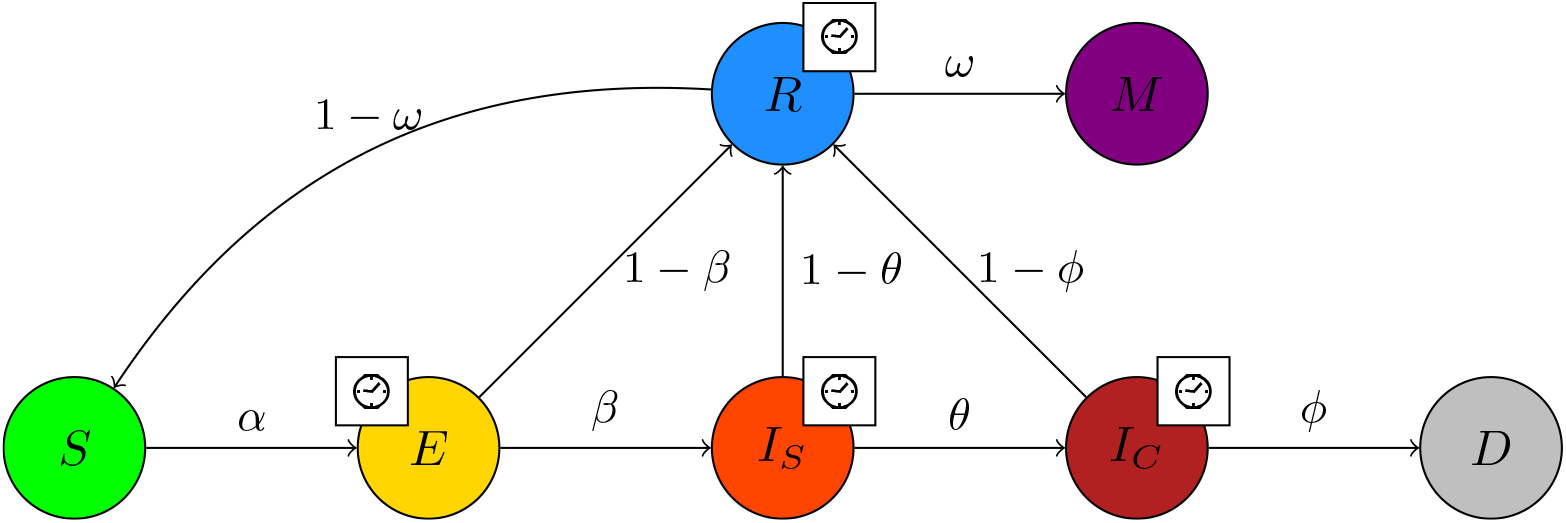
Transmission dynamics of the infectious disease at individual level in INFEKTA. The infectious disease states are, Susceptible (*S*), Exposed (*E*), Seriously-Infected (*I*_*S*_), Critically-Infected (*I*_*C*_), Recovered (*R*), Immune (*M*) and Dead (*D*). Probabilities are individual based and can be defined in terms of current location, age, gender, and so on. Symbol 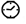, on an state *X*, indicates that an individual must stay some period of time 𝒯_*X*_ at such state *X* before being able to change to other state.

Since rates are defined at the individual level, these rates can be defined by taking into consideration, for example, rates at the population level (obtained from a compartmental model), age, gender and co-morbidity presented in the individual. Remember that those rates are not defined at some time scale (as in compartmental models) but define the rule determining changes in health states of individuals being close enough for interacting at the infectious disease transmission level or after some period of time being in some state. In this way, an individual can change with probability *α* from *S* state to state *E* if close enough to an infected (*E, I*_*S*_, and *I*_*C*_) individual and will change from state *I*_*S*_ to state *I*_*C*_ if has been at state *I*_*S*_ with probability *θ* if has been on state *I*_*S*_ a period of time 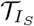.

- *α*: is the transmission rate and incorporates the encounter rate between susceptible and infectious individuals together with the probability of transmission.
- *β*: is the rate at which individuals move from the exposed (*E*) to the Seriously-Infected state (*I*_*S*_). It’s reciprocal (1*/β*) is the average latent (exposed) period.
- *θ*: is the rate at which individuals move from the Seriously-Infected (*E*) to the Critically-Infected state (*I*_*S*_). Its reciprocal (1*/θ*) is the average critical period.
- *ϕ*: is the death rate.
- *ω*: is the immune rate that incorporates the probability of becoming immune.
- 𝒯_*E*_: Time an individual will be at the Exposed state (*E*) before changing to the Recovered (*R*) or Seriously Infected (*I*_*S*_) states.
- 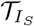 Time an individual will be at the Seriously Infected state (*I*_*S*_) before changing to the Recovered (*R*) or Critically Infected (*I*_*C*_) states.
- 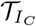 Time an individual will be at the Critically Infected (*I*_*C*_) before changing to the Recovered (*R*) or Dead (*D*) states.
- 𝒯_*R*_: Time an individual will be at the Recovered state (*R*) before changing to the Susceptible (*S*) or Immune (*M*) states.

INFEKTA can consider that two individuals were close enough for interacting at the transmission of the infectious disease if they were at the same place (home, interest place or public transportation station) at the same time or if they were close enough (in the Euclidean space) while moving. In order to simplify this checking process, it is possible to consider that an individual just visited its home, final interest place, and both the initial and final PTSs when using the public transportation system.

### Social Separation Policy

The social separation policy is described in INFEKTA as a finite sequence of rules, each rule having **i) Start Time**: an initial day for applying the social separation policy rule; **ii) End Time**: final day for ending the social separation policy rule; **iii) Level**: indicates the kind of restriction applied to the mobility of persons and accesses to places, and **iv) Enforce**: defines the specific mobility and access restrictions of the social separation policy.

### Modeling the Transmission Dynamics of the COVID-19 in Bogotá - Colombia

INFEKTA is used for modeling the Transmission dynamic of the COVID-19 in Bogotá city, the largest and crowded city in Colombia. Bogotá is the capital city of Colombia, its urban perimeter population is 7.412.566, is composed by 20 *districts*, and its massive public transportation system is called *Transmilenio* (**TM**). TM is a bus-based system, which has 143 stations and moves near to 2.500.000 citizens every day.

#### Virtual Space Setup

Geographical information of Bogotá is used as the Euclidean space where the moving and interaction complex network is defined. Each one of the TM stations is located and added to the complex network according to the real TM system [27]. Also, the airport and the regional bus terminal are located and connected to the nearest TM station.

Demographic information from nineteen districts of Bogotá (only urban districts) is used for generating in the Euclidean space interest places (Workplaces (W), markets (M), and schools (S)), homes (H), and people(P). Places are generated, in each one of the districts, following a 2D multivariate normal distribution *N* ∼ (*µ*, Σ) (*µ* is the geographic center of the district and Σ is the co-variance matrix defined by the points determining the perimeter of the district). The number of places in each district was generated based on the population density of each district according to the projections to 2015 [28]. Table 1 shows the amount of data generated for each type of place and for people, also the number of TM stations (Bus), and terminal transportation that we use in the simulation, and Table 2 shows a detailed information of the number of interest places generated by district.

**Table 1.** Data used in the simulation. (*) real places.

| Agent | Instance | Type | Amount |
| --- | --- | --- | --- |
| Place | Home (H) | Home (H) | 309 |
|  | Public Transportation Station (PTS) | Bus (B)* | 143 |
|  | Interest place (IP) | Workplace (W) | 129 |
|  |  | School (S) | 68 |
|  |  | Market (M) | 110 |
|  |  | Terminal (T)* | 2 |
| Individual | Individual | People (P) | 1001 |

**Table 2.** Number of interest places generated by districts.

| DISTRICT | Amount of places |  |  |  |
| --- | --- | --- | --- | --- |
|  | Homes(H) | Schools(S) | Markets(M) | Workplaces(W) |
| (01) Usaquén | 19 | 4 | 7 | 8 |
| (02) Chapinero | 6 | 2 | 2 | 3 |
| (03) Santa Fe | 5 | 1 | 2 | 2 |
| (04) San Cristóbal | 16 | 4 | 6 | 7 |
| (05) Usme | 17 | 4 | 6 | 7 |
| (06) Tunjuelito | 8 | 2 | 3 | 4 |
| (07) Bosa | 25 | 5 | 9 | 10 |
| (08) Kennedy | 41 | 9 | 14 | 17 |
| (09) Fontibón | 15 | 3 | 5 | 6 |
| (10) Engativá | 34 | 7 | 12 | 14 |
| (11) Suba | 45 | 9 | 15 | 18 |
| (12) Barrios Unidos | 10 | 2 | 4 | 4 |
| (13) Teusaquillo | 6 | 2 | 2 | 3 |
| (14) Los Mártires | 4 | 1 | 2 | 2 |
| (15) Antonio Nariño | 5 | 1 | 2 | 2 |
| (16) Puente Aranda | 10 | 2 | 4 | 4 |
| (17) La Candelaria | 1 | 1 | 1 | 1 |
| (18) Rafael Uribe | 15 | 3 | 5 | 6 |
| (19) Ciudad Bolívar | 27 | 6 | 9 | 11 |
| **TOTAL** | **309** | **68** | **110** | **129** |

Figure 2 shows the virtual Euclidean map of Bogotá with the georeferenced real and simulated places; also, the figure shows the associated complex network of connected places (nodes are places and edges are routes between places), the graph was drawn with Gephi [29].

**Fig 2.**
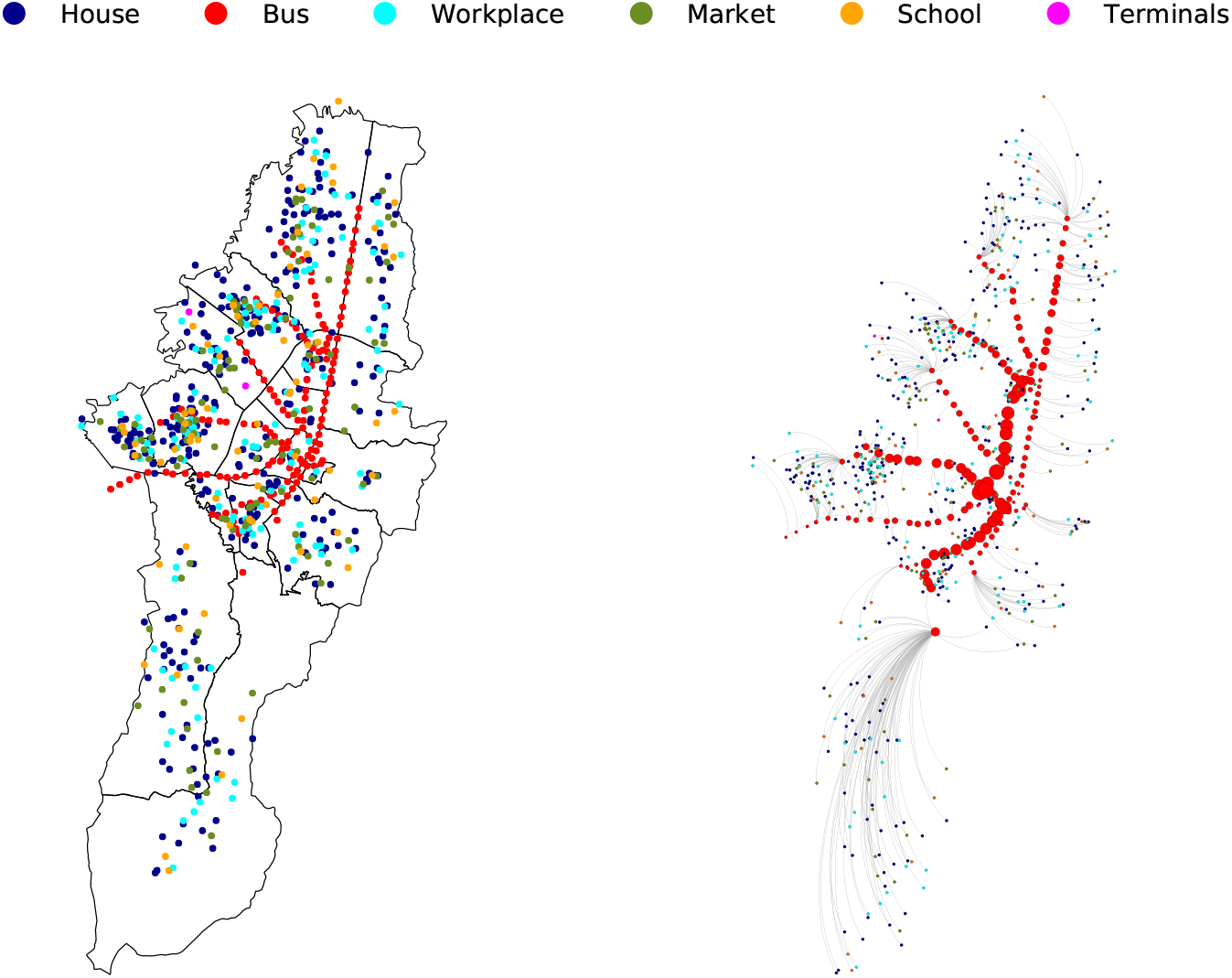
Bogotá with georeferenced places (left) and Bogotá complex network (right).

#### Individuals setup

An heterogeneous (varying gender, age, district and home) group of one thousand one individuals (1001) is generated using a stratified sampling based on the demographic information of the city for each district according to the projections to 2015 [28]. An individual is classified, according to her/his age, as: *Child* = [0-12], *Adolescence* = [13-18], *Adult* = [19-49], *Senior* = [50-69], and *older* = 70+. Table 3 shows the total demographic information of virtual people.

**Table 3.** Demographic information of virtual people. District (D), Total(T), Male(M), Female(F).

| D | TOTAL |  |  | Child<br>[0-12] |  |  | Adolescence<br>[13-18] |  |  | Adult<br>[19-49] |  |  | Senior<br>[50-69] |  |  | older<br>70+ |  |  |
| --- | --- | --- | --- | --- | --- | --- | --- | --- | --- | --- | --- | --- | --- | --- | --- | --- | --- | --- |
|  | T | M | F | T | M | F | T | M | F | T | M | F | T | M | F | T | M | F |
| (01) | 63 | 35 | 28 | 13 | 4 | 9 | 5 | 4 | 1 | 32 | 21 | 11 | 11 | 5 | 6 | 2 | 1 | 1 |
| (02) | 17 | 7 | 10 | 1 | 0 | 1 | 1 | 0 | 1 | 9 | 5 | 4 | 4 | 2 | 2 | 2 | 0 | 2 |
| (03) | 14 | 9 | 5 | 1 | 1 | 0 | 1 | 0 | 1 | 8 | 5 | 3 | 4 | 3 | 1 | 0 | 0 | 0 |
| (04) | 52 | 32 | 20 | 13 | 9 | 4 | 3 | 1 | 2 | 20 | 13 | 7 | 14 | 8 | 6 | 2 | 1 | 1 |
| (05) | 55 | 33 | 22 | 21 | 11 | 10 | 4 | 3 | 1 | 21 | 13 | 8 | 5 | 2 | 3 | 4 | 4 | 0 |
| (06) | 25 | 14 | 11 | 5 | 4 | 1 | 3 | 2 | 1 | 10 | 4 | 6 | 3 | 2 | 1 | 4 | 2 | 2 |
| (07) | 82 | 45 | 37 | 19 | 7 | 12 | 7 | 4 | 3 | 42 | 23 | 19 | 12 | 9 | 3 | 2 | 2 | 0 |
| (08) | 137 | 82 | 55 | 26 | 12 | 14 | 11 | 6 | 5 | 66 | 45 | 21 | 27 | 14 | 13 | 7 | 5 | 2 |
| (09) | 48 | 24 | 24 | 9 | 5 | 4 | 3 | 0 | 3 | 22 | 12 | 10 | 13 | 6 | 7 | 1 | 1 | 0 |
| (10) | 112 | 50 | 62 | 16 | 5 | 11 | 12 | 5 | 7 | 51 | 23 | 28 | 25 | 13 | 12 | 8 | 4 | 4 |
| (11) | 150 | 73 | 77 | 35 | 13 | 22 | 14 | 7 | 7 | 74 | 37 | 37 | 23 | 14 | 9 | 4 | 2 | 2 |
| (12) | 30 | 13 | 17 | 5 | 3 | 2 | 2 | 1 | 1 | 13 | 6 | 7 | 9 | 2 | 7 | 1 | 1 | 0 |
| (13) | 19 | 8 | 11 | 3 | 2 | 1 | 2 | 0 | 2 | 10 | 5 | 5 | 2 | 0 | 2 | 2 | 1 | 1 |
| (14) | 12 | 5 | 7 | 3 | 1 | 2 | 1 | 0 | 1 | 3 | 1 | 2 | 5 | 3 | 2 | 0 | 0 | 0 |
| (15) | 13 | 6 | 7 | 3 | 2 | 1 | 1 | 0 | 1 | 4 | 2 | 2 | 4 | 2 | 2 | 1 | 0 | 1 |
| (16) | 33 | 14 | 19 | 6 | 2 | 4 | 5 | 1 | 4 | 14 | 6 | 8 | 1 | 1 | 0 | 7 | 4 | 3 |
| (17) | 3 | 2 | 1 | 0 | 0 | 0 | 0 | 0 | 0 | 3 | 2 | 1 | 0 | 0 | 0 | 0 | 0 | 0 |
| (18) | 48 | 29 | 19 | 11 | 8 | 3 | 6 | 1 | 5 | 24 | 14 | 10 | 5 | 4 | 1 | 2 | 2 | 0 |
| (19) | 88 | 43 | 45 | 18 | 8 | 10 | 7 | 3 | 4 | 48 | 23 | 25 | 10 | 6 | 4 | 5 | 3 | 2 |
|  | 1001 | 524 | 477 | 208 | 97 | 111 | 88 | 38 | 50 | 474 | 260 | 214 | 177 | 96 | 81 | 54 | 33 | 21 |

Also, a sequence of activities was assigned randomly to each individual to define a diary routine. This was done according to the person’s age and the hour of the day. For example, some agents *Adolescence* go to school, and some agents *adult* go to work. Some agents may move using the PTI system and some others while going directly to its destination place. The route an individual takes is defined according to the complex network. Figure 3 shows three examples of different routines (paths over the graph) for the individuals.

**Fig 3.**
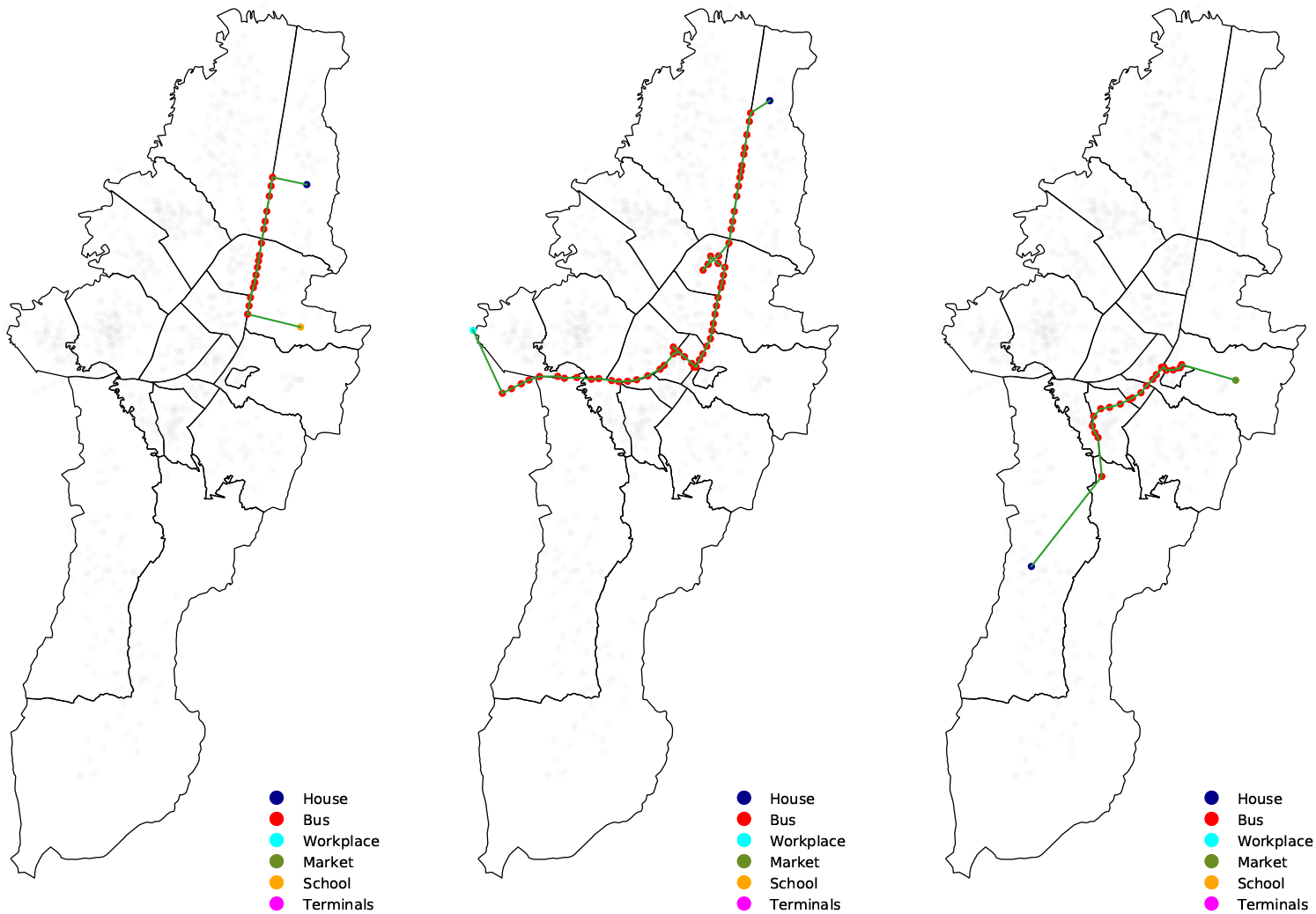
Example routines (paths) carried by individuals [Age, Gender, IP, District]. Individual 5 (left): [Child, F, School, 01]; Individual 55 (middle): [Senior, F, Workplace, 01]; Individual 999 (right): [Older, M, Market, 19].

The explicit impact level of age and medical preconditions on the state of the COVID-19 dynamic is not included in this preliminary modeling. We wrapped them in the transition rates and allow modelers to change and play with different rates. Therefore, we set the initial values of these rates as shown in Table 4.

**Table 4.** Parameters of INFEKTA and their estimations for COVID-19.

| Symbol | Description | COVID-19 estimations |
| --- | --- | --- |
| $\alpha$ | Probability of $S \rightarrow E$ | 0.1 |
| $\beta$ | Probability of $E \rightarrow I_S$ | 0.4 |
| $\theta$ | Probability of $I_S \rightarrow I_C$ | 0.4 |
| $\phi$ | Probability of $I_C \rightarrow D$ | 0.1 |
| $\omega$ | Probability of $R \rightarrow M$ | 0.1 |
| $\mathcal{T}_E$ | Time at $E$ | 5 days |
| $\mathcal{T}_{I_S}$ | Time at $I_S$ | 5 days |
| $\mathcal{T}_{I_C}$ | Time at $I_C$ | 5 days |
| $\mathcal{T}_R$ | Time at $R$ | 5 days |

#### Social Separation Rule Setting

The level attribute of the social separation rule for the COVID-19 in the virtual Bogotá city is defined as follows:

- None: No restrictions to the mobility neither to access to places.
- Soft: Few places are restricted (depending on the type, capacity, etc). i.e., randomly close *U* ∼ (*size*(*places*) ∗ 0.3, *size*(*places*) ∗ 0.5).
- Medium: Many places and few stations are restricted (depending on the type, capacity, etc). Some type of individuals is restricted to stay at home (except those with the required mobility level). i.e., randomly close *U* ∼ (*size*(*places*) ∗ 0.5, *size*(*places*) ∗ 0.7).
- Extreme: Few places are accessible to persons while few stations are restricted. Almost every individual is restricted to stay at home (except those with the required mobility level). i.e., randomly close *U* ∼ (*size*(*places*) ∗ 0.7, *size*(*places*) ∗ 0.9).
- Total: Scare places are accessible to persons, and almost all stations are not accessible. Individuals are restricted to stay at home (except those with the required mobility level). i.e., randomly close *U* ∼ (*size*(*places*) ∗ 0.9, *size*(*places*)).

## Results

The INFEKTA implementation of the COVID-19 propagation model for Bogotá city is implemented using the simulation modeling software tool *AnyLogic*, see S1 Simulator. Also, the methodology (Data preprocessing; places, population, and routes assignation; network creation) is available in a Github repository, see S2 Repository.

Figure 4 shows the initial configuration of virtual individuals in the simulation, represented as small points. All of them are initially assigned to homes, and almost all of them are in the Susceptible state (*S*) (green points). Just ten (10) individuals are considered at state Exposed (*E*).

**Fig 4.**
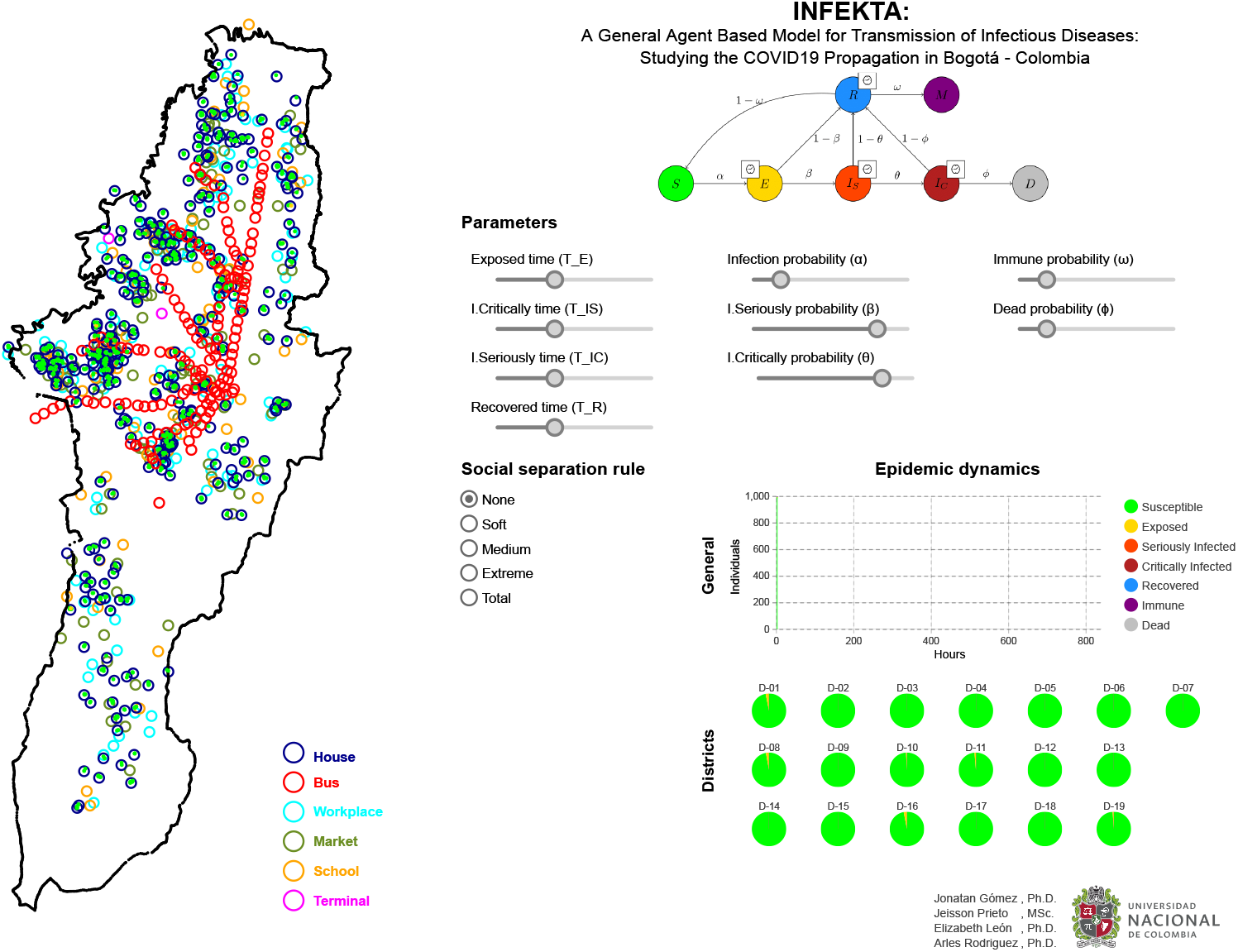
Initial configuration of INFEKTA. The simulator presents the dynamic of the Individuals (agents) in Bogotá, the parameters of INFEKTA, the social separat**P**i**a**o**us**n**ed** rules that can be applied, and the Epidemic dynamics (agents state per hour).

Figure 5 shows the state of the COVID-19 dynamics (propagation of the virus) at three different simulation time steps. Figure 5 (left) shows such dynamics after some simulated minutes of starting the simulation. Notice that, individuals are moving (mainly using Transmilenio) according to their assigned routines. Figure 5 (middle) shows the COVID-19 dynamics after 400 simulated hours. At this time, all individuals have been Exposed (yellow points), meaning that some individuals are seriously infected (red points), critically infected (dark red points), recovered (blue points), and few of them become immune (purple points). Figure 5 (right) shows the COVID-19 dynamics after 800 simulated hours. Notice that, the virus has already peaked once and the first wave is over. The propagation is now stable and 20% of the population is now immune to the virus. As expected, the number of exposed, seriously and critically infected cases grows exponentially when a social separation policy is not enforced. Clearly, the transmission dynamics of the infectious disease (in this case COVID19) emerge from the local interaction of the individuals.

**Fig 5.**
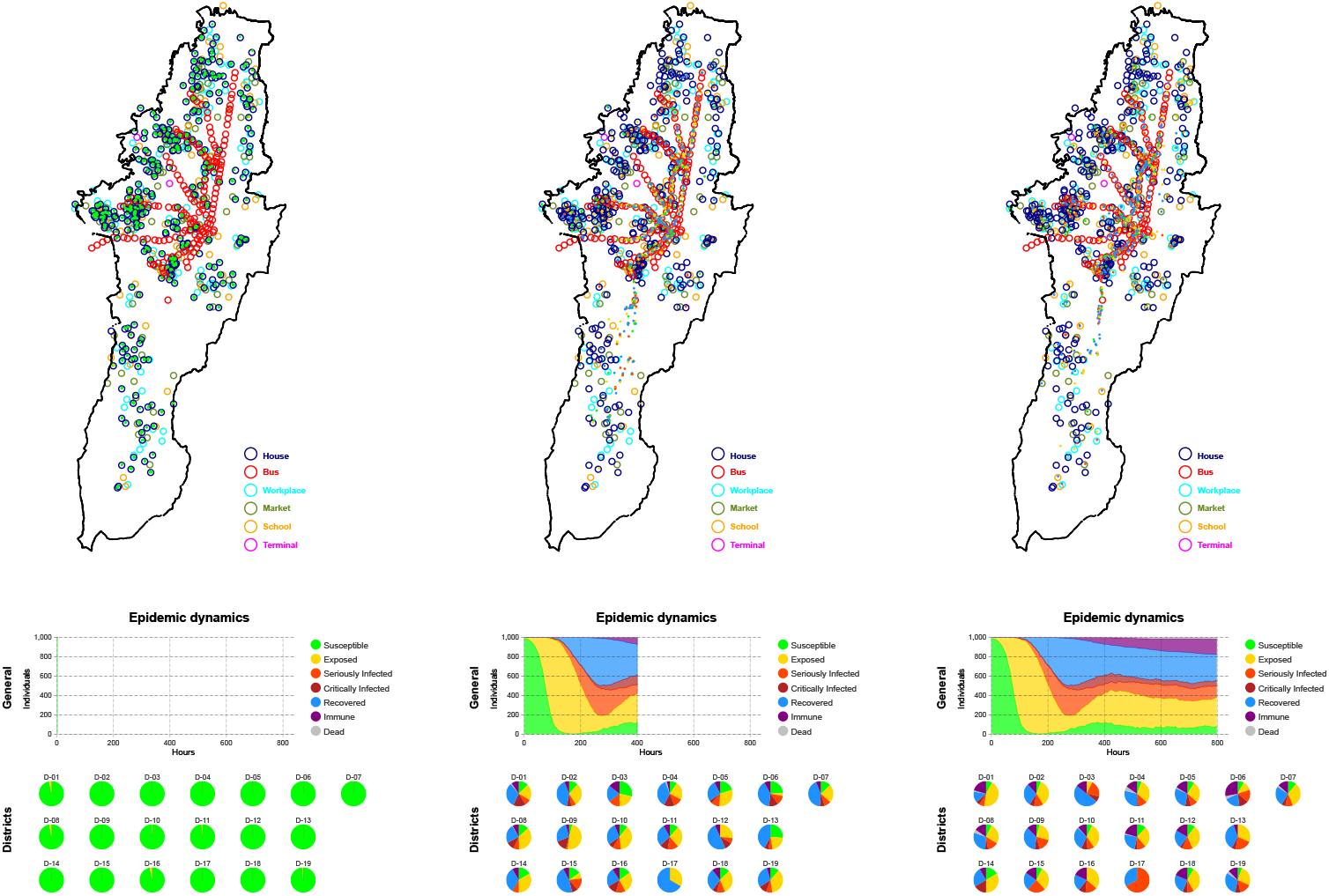
Evolution of the epidemic dynamics of INFEKTA. Initial configuration (left), 400 hours (middle), and 800 hours (right).

We analyze the sensitivity of some parameters to check the robustness of the model. Sensitivity to infection rate is shown in Figure 6. Notice that by increasing or decreasing the infection disease rate (Figure 6 (left)), the peak of the transmission dynamics is reached sooner or later on time. When low infection diseases rates, the number of cases is also low, reducing the impact on the economy but a new peak is likely to appear after that. On the other hand, for high infection disease rates (Figure 6 (right)) the peak is reached in an early stage, and around half of the population is on one of the infected states (Exposed, Seriously-Infected, Critically-Infected).

**Fig 6.**
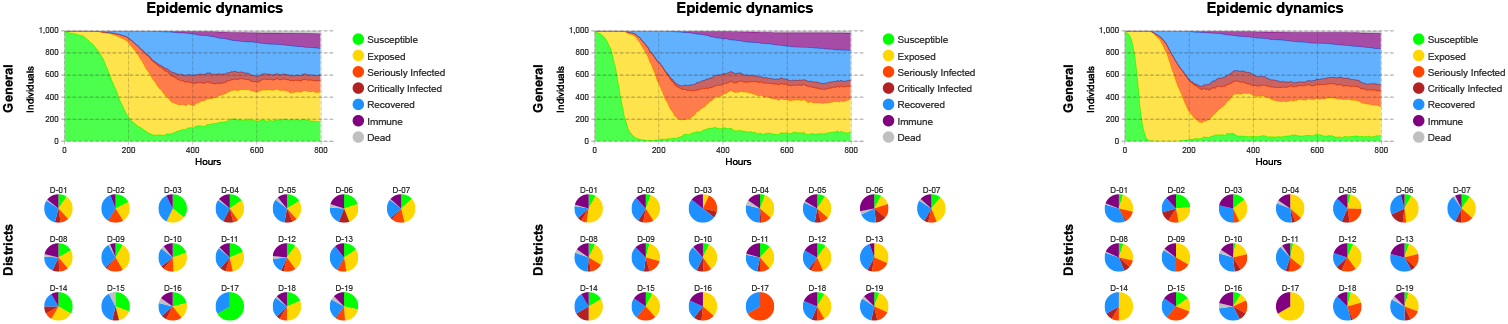
Sensitive analysis for the infection probability. *α. α* = 0.05 (left), *α* = 0.1 (middle), *α* = 0.2 (right).

Sensitive to recovery time (𝒯_*R*_) is shown in Figure 7. If the recovery time is reduced to 2 days (Figure 7 (left)) there is not a significant difference in the epidemic behavior respect to the predefined 5 days. However, if recovery time is increased to 10 days (Figure 7 (right)) the transmission disease dynamics show a second peak after 500 hours (approx 21 days).

**Fig 7.**
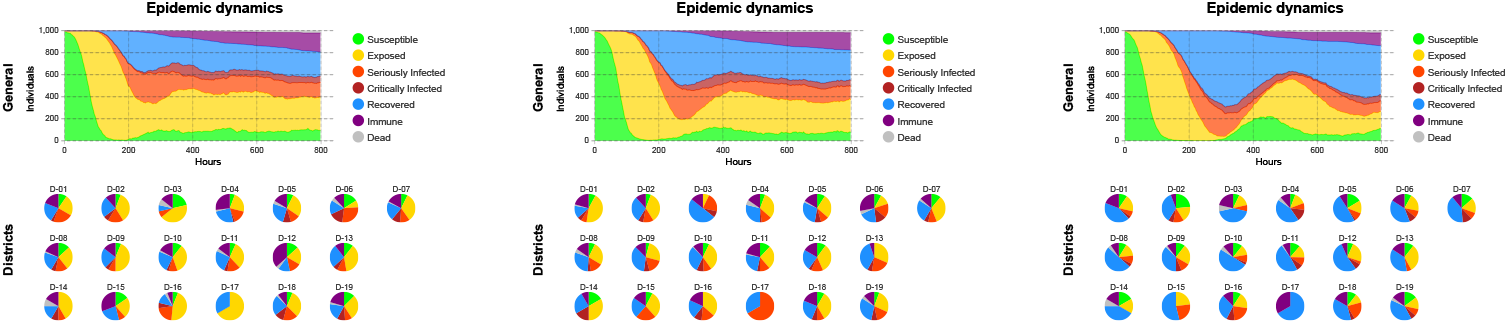
Sensitive analysis for the Recovery Time. 𝒯_*R*_. 𝒯_*R*_ = 2 days (left), 𝒯_*R*_ = 5 days (middle), 𝒯_*R*_ = 10 days (right).

Also, we try different scenarios where each one of the social separation policies is enforced just after 800 simulation hours, see Figure 8. As can be noticed, its evident how social separation rules help to mitigate the exponential growth on the transmission disease dynamics (COVID19), reducing the number of infectious cases (Exposed, Seriously-Infected, and Critically-Infected). Interestingly, when the extreme social separation rule (access to approximately 75% of interest places is restricted) the transmission disease dynamic displays similar behavior to the one when a total social separation rule (access to approximately 95% of interest places is restricted) is enforced.

**Fig 8.**
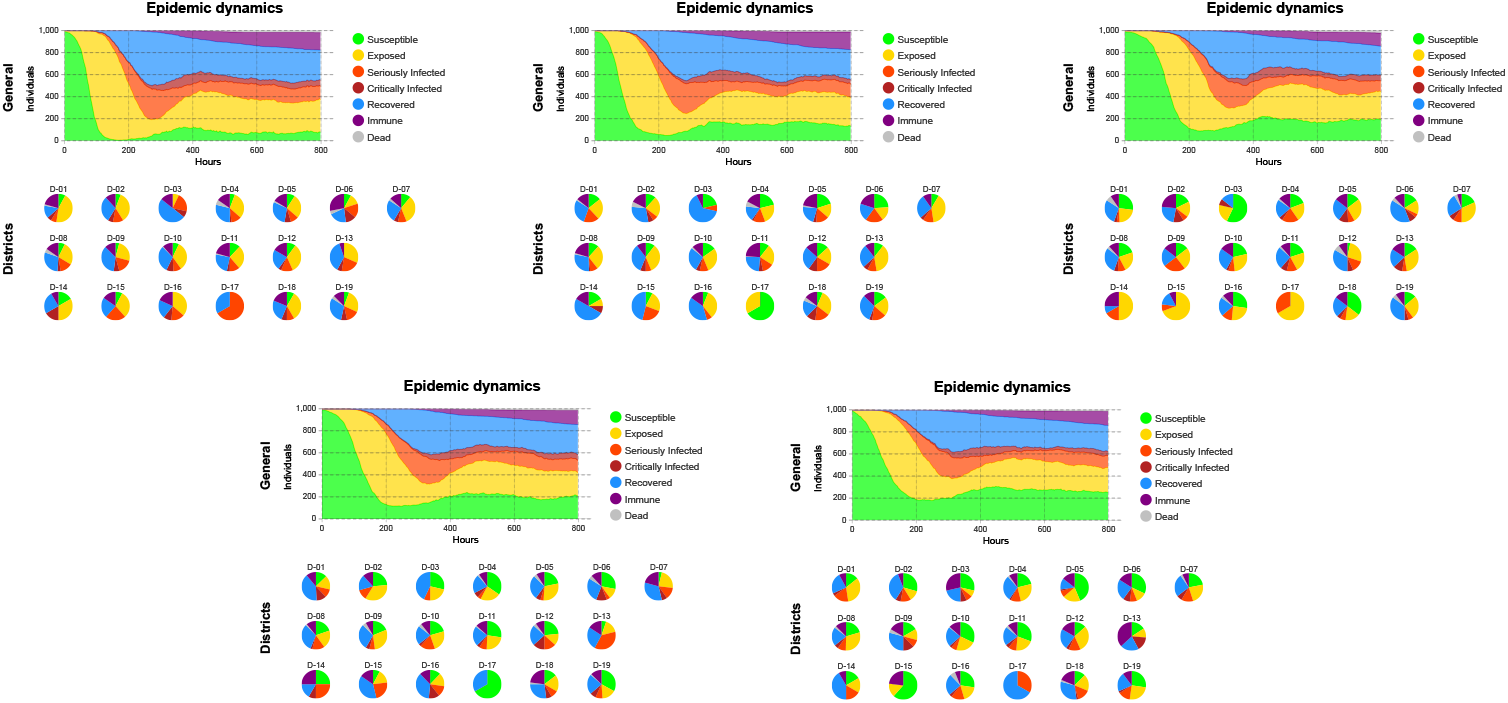
Social Separation after 800 hours and initialized in hour 0. None (top-left), Soft (top-middle), Medium (top-right), Extreme (bottom-left), and Total (bottom-right).

## Conclusions and Future Work

Modeling the Transmission dynamic of an infectious disease such as the COVID-19 is not an easy task due to its highly complex nature. When using an agent-based model, several different characteristics can be modeled, for example, the demographic information of the population being studied, the set of places and the mobility of agents in the city or town under consideration, social separation rules that may be enforced, and the special characteristics of the infectious disease being modeled. INFEKTA is a general agent-based model that allows researches to combine and study all of those characteristics.

Our preliminary results modeling the transmission dynamics of the coronavirus COVID-19 in Bogotá city, the largest and crowded city in Colombia, indicate that INFEKTA may be a valuable asset for researchers and public health decision-makers for projecting future scenarios when applying different social separation policy rules and controlling the expansion of an infectious disease. Although we are doing a rough and no so real approximation of the transmission dynamics of the COVID19, we are able to obtain similar behaviors, in our preliminary experiments, to those reported for the COVID19 in the real world. Therefore, INFEKTA may be able to provide more accurate results if its parameters are set to real ones: disease transmission rates, virus, incubation periods, comorbidity, houses, interest places, routines, population size (close to nine millions of virtual individuals for the Bogotá city).

Our future work will concentrate on studying the transmission of COVID-19 in Bogotá city by considering different scenarios of social separation rules and by using more realistic information about: i) Relation between personal information and propagation rates of the COVID-19, ii) Places and routes, iii) Population size, and iv) Age and Medical preconditions.

## Data Availability

No medical data from external sources is used in this paper. Demographic data of the Bogotá city is available on Colombian government sites. Transmilenio data is available at http://www.transmilenio.gov.co

https://cloud.anylogic.com/model/9806d370-f0a8-48d1-b3e1-3537721b39ba

https://github.com/japrietov/INFEKTA

## Supporting information

**S1 Simulator. INFEKTA Anylogic Simulator**. A hands-on version where anybody can try to change some of the parameters is available at INFEKTA anylogic.

**S2 Repository. INFEKTA repository**. A repository containing the source code of the simulator and a technical report explaining the modeling methodology is available at INFEKTA github.

## Footnotes

1 in the 2D Euclidean space.

